# Practical Strategies for Extreme Missing Data Imputation in Dementia Diagnosis

**DOI:** 10.1101/2020.07.13.20146118

**Authors:** Niamh McCombe, Shuo Liu, Xuemei Ding, Girijesh Prasad, Magda Bucholc, David P. Finn, Stephen Todd, Paula L. McClean, KongFatt Wong-Lin, Alzheimer’s Disease Neuroimaging Initiative (ADNI)

## Abstract

Accurate computational models for clinical decision support systems require clean and reliable data but, in clinical practice, data are often incomplete. Hence, missing data could arise not only from training datasets but also test datasets which could consist of a single undiagnosed case, an individual. This work addresses the problem of extreme missingness in both training and test data by evaluating multiple imputation and classification workflows based on both diagnostic classification accuracy and computational cost. Extreme missingness is defined as having ∼50% of the total data missing in more than half the data features. In particular, we focus on dementia diagnosis due to long time delays, high variability, high attrition rates and lack of practical data imputation strategies in its diagnostic pathway. We identified and replicated the extreme missingness structure of data from a real-world memory clinic on a larger open dataset, with the original complete data acting as ground truth. Overall, we found that computational cost, but not accuracy, varies widely for various imputation and classification approaches. Particularly, we found that iterative imputation on the training dataset combined with a reduced-feature classification model provides the best approach, in terms of speed and accuracy. Taken together, this work has elucidated important factors to be considered when developing a predictive model for a dementia diagnostic support system.

## I. Introduction

The issue of missing data is one of the most ubiquitous concerns in data science [1]. This is particularly the case in clinical and medical data, which frequently has many missing values [2]–[4] (see Fig. 1a for a real-world, routine (i.e. not clinical trial) Alzheimer’s disease (AD) dataset). In recent years, there has been increased effort to assure data quality and reusability, and to automate the processes of discovering and analysing data by publishing data annotations and analytical workflows [5], [6].

**Fig. 1.**
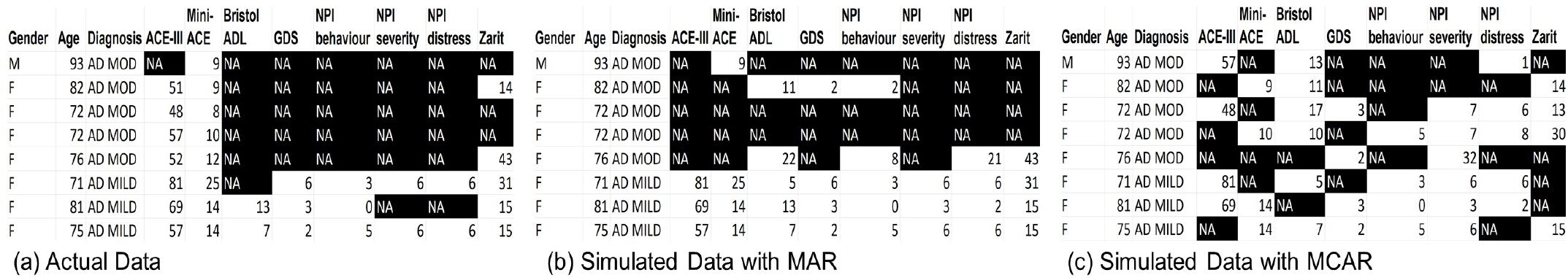
Sample Alzheimer’s disease (AD) dataset from a memory clinic and its breakdown of data missingness. (a) Actual sample data. Rows: patients; columns: diagnosis category (AD MILD or AD MOD for mild or moderate AD, respectively), the various cognitive and functional assessments, Gender and Age. Black cells with “NA” label: missing data. (b-c) Simulated data with missingness correlated with diagnosis (Missing at Random, MAR) (b), and uncorrelated with any variable (Missing Completely at Random, MCAR) (c).

A key clinical application of data science is in the development and use of computerized decision support systems (CDSS), which can enhance consistency, objectivity and standardization [6]–[8] In developing a clinical diagnostic model for use in a CDSS, large training dataset is typically used to build a classification model, while test dataset is used to verify model accuracy [9]. Generally, the training and test datasets must be complete, with no missing values for any variables. In cases of extreme missingness, which we define as having ∼50% of the total data missing in more than half the data features, which often occurs in real-world routine clinical data records, it may not be practical or possible to acquire the missing data to improve data modelling. Hence, computational models must incorporate a strategy (method or combination of methods) for handling missing data as part of their analytical workflow.

Current strategies for handling missing data include: (i) attempting to acquire missing data at additional expense, e.g. performing an assessment which was previously not conducted; (ii) complete-case analysis, in which any row with a missing value is dropped from analysis; (iii) data imputation, in which missing values are replaced with an estimated value; (iv) missing-indicator methods, in which missing values are marked as missing and then incorporated in the training dataset; and (v) various strategies in which missing data is tackled directly in the analysis without an intermediate imputation step [10]–[12]. The latter includes maximum-likelihood methods [13], classifiers which can account for the uncertainty caused by missing data such as the naïve credal classifier [14], and tree-based classifiers which use the surrogate split method [15].

Data imputation strategies can further be divided into single imputation methods, in which a single estimate for the missing data is generated, and multiple imputation methods, which generate multiple estimates for each missing value and therefore will produce multiple imputed datasets for further analysis [2], [16]. Another crucial distinction is between supervised data imputation methods, where the class label is known, and unsupervised methods, which operate in the absence of a class label [17]. It is also useful to highlight that many commonly used imputation methods are iterative imputation methods which impute the entire dataset repeatedly until an optimum is reached e.g. [18], [19].

The appropriate strategy for dealing with missing data will depend to some extent on the type of missingness. Missing data is often categorized into three types: missing at random (MAR); missing completely at random (MCAR); and missing not at random (MNAR) [20]. In the case of MAR, the probability that data is missing depends upon the variables observed within the dataset. Fig. 1b shows a simulated sample AD dataset in which cognitive testing variables are more likely to be missing in more severe AD cases due to the difficulty of performing cognitive assessments on such patients. MCAR can be understood as a special case of MAR – in this case, the probability of missingness is independent of all variables in the dataset. An example would be someone being late for a medical appointment because of a traffic jam so there would be insufficient time to complete all of their cognitive assessments (see Fig. 1c for a simulated sample example of MCAR). MNAR is the case where the probability of missingness depends on a variable which is in itself missing; this is the most complex case to handle. An example of this might be a survey on income, in which people with a very low or very high income refuse to report their income [2]. MNAR type missingness is also very common in longitudinal data e.g. a clinical dataset where disease progression may lead to subjects dropping out of the study [21], [22]. Importantly, longitudinal studies on cognitive decline have high attrition rates (e.g. [23]–[25]).

In practice, clinical data tends to have MAR type missingness [2]. However the probability of missingness in clinical data is often dependent on the outcome variable, as illness/disease severity may impact opportunities for data gathering [26]. In longitudinal data, this may be MNAR type missingness, such as the case where a study participant may not be able to undergo a specific assessment or be part of a follow up study due to an increase in disease severity. The correlation between missingness and disease severity holds true in dementia data, as shown in [21].

Various studies have evaluated different imputation methods for replacing missing values in clinical data [2], [16], [27]–[30]. The most effective methods are found to be multivariate, iterative methods such as Multiple Imputation by Chained Equations (MICE) [29] fuzzy k-means [16], [27], Bayesian Principal Component Analysis [27] and missForest [18], and more recently, unsupervised neural network’s autoencoders [31]. However, most studies are focused on handling missingness in the training dataset, despite the fact that the test dataset can have missing values. For example, the diagnosis of a patient may involve unknown data variables from that patient (Fig. 2).

**Fig. 2.**
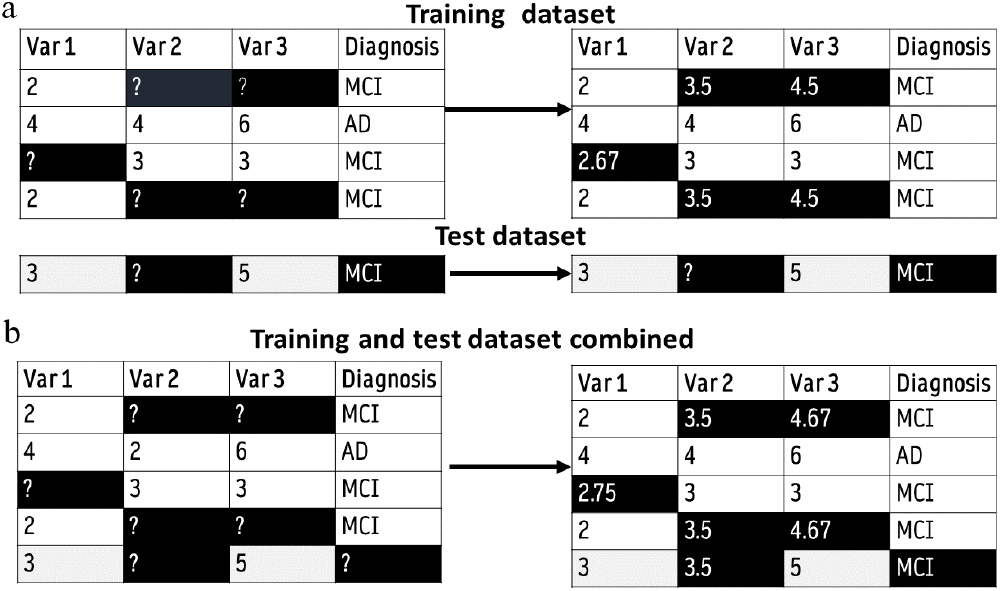
Iterative imputation with single-row test dataset of a toy example. Iterative imputation begins with mean imputation. (a) It is impossible to separately impute training and test datasets when test dataset is very small. (b) The training and test datasets are imputed together. Thus, the computational time to impute test dataset is the same as imputing the entire dataset. Note: To test classification accuracy, the class variable for the test dataset must be removed and imputed, to avoid ‘double-dipping’.

The case of missing values in the test dataset during classification was addressed in [32], which also notes the dearth of literature on this issue. Specifically, [32] delineated four different strategies for handling the situation of missing values in the test data: (i) discarding objects with missing values; (ii) acquiring the missing value through manual follow-up; (iii) data imputation; or (iv) using a reduced-feature classification model built with variables which are not missing in the test dataset, and concluding that reduced-feature methods provide an under-utilised and efficient solution to the problem of missing values in the test dataset. Another study evaluated strategies for missing values in the test data in the context of a tree-based classifier and for eight different missing data patterns, using simple datasets with a binary response variable [33]. The conclusion was that a missing-indicator method was the most useful where missingness is related to the response variable. A later study [34] directly addressed the problem of missing values in the test clinical dataset, using k-nearest neighbors (k-NN) imputation method [35] to impute the dataset before testing the impact on classification accuracy, finding that even when 25% of the values are missing it is possible to achieve good classification accuracy.

It is clear that the above studies for handling missing test data are limited. Specifically, [32] and [33] had yet to test their methods on real-world clinical data, and did not discuss the issue of missing training data, while the workflow in [34] appeared to have training and test datasets imputed together. In particular, iterative imputation methods of handling missing data may be unsuitable to apply to small test data in real time (Fig. 2), potentially limiting the usefulness of such methods in a clinical decision-making context. Additionally, very little missing data literature deals with extreme missingness. Importantly, there is no literature on missing data that deals with the specific prerequisites that are likely to be present in a clinical decision-making setting, notably: (i) when a patient is being diagnosed (corresponding to classification in machine learning models), it is likely that there will be significant missing data related to that patient (missing test data); (ii) patients are diagnosed one at a time by clinicians, corresponding to leave-one-out cross validation (LOOCV) condition for testing machine-learning models within a CDSS (e.g. [36], [37]), and (iii) imputation and classification of the test dataset must be performed within a reasonable timeframe for efficient and timely diagnosis.

In this work, we investigate strategies for handling extreme missing data which takes these constraints into consideration, with missing data patterns that resemble those from real-world, routine clinical data. We focus on the diagnosis of dementia, particularly Alzheimer’s disease (AD), due to AD being the most common form of dementia, and AD’s long time delays and high variability in its diagnostic pathway [22]. Additionally, there is a substantial scarcity of practical data imputation strategies for dementia diagnosis (e.g. [22], [38]–[44]).

## II. Methods

### A. Data Description

#### 1) Clinical Dataset to Extract Missing Data Characteristics

Anonymous clinical data were extracted from Altnagelvin Area Hospital’s Memory Assessment Service (WHSCT) in the form of a CSV file. Ethics approval for this was obtained from the Office for Research Ethics Committee Northern Ireland (ORECNI, HSC REC B reference: 17/NI/0142; IRAS project ID: 230077). This data was used to determine the type of missingness in a real-world, routine clinical dataset to reproduce in the ADNI dataset. A sample of the dataset is shown in Fig. 1A. There were 189 rows in total, each representing a patient. Cells with missing values are shown in black in the diagram. Features included 7 different Cognitive and Functional Assessment (CFA) scores as well as Gender, Age and text-based Diagnosis information. AD diagnosis was manually categorized into two classes, 85 AD MILD (mild AD) and 104 AD MOD (moderate AD). Other diagnostic categories, including non-AD dementia subtypes, were discarded due to lack of ordinality or their small sizes. In our previous work, we showed that CFAs are among the most predictive features for classifying AD severity [37], [45]. For the current clinical dataset, the CFAs included Addenbrooke’s Cognitive Examination (ACE-III) and the Mini-ACE [46], the Bristol Activities of Daily Living Scale [47], the Geriatric Depression Scale [48], the NPI-Q behavioral, distress and severity measurements [49], and the Zarit Caregiver Burden [50]. Hence, this study focuses on CFA features. The extracted missingness structure of this dataset was replicated in a complete open source dataset, as described below.

#### 2) ADNI Dataset

The data for evaluating the missing data strategies was extracted from the ADNIMERGE table [51] from the Alzheimer’s Disease Neuroimaging Initiative (ADNI) merge R package, which amalgamates several key tables from the ADNI open source dementia data (adni.loni.usc.edu). The ADNI open database included clinical and neuropsychological assessments with diagnosis labelled as healthy, mild cognitive impairment (MCI) and early AD. It should be noted that the MCI group may include prodromal stage of AD, and individuals who will not progress to AD. After feature selection (see Section II.B.1) was applied to ADNIMERGE CFA variables, we had 8 CFA variables in the dataset (see Table I). We also included Gender and Age in our analysis, mirroring the routine clinical dataset, and the CFA MMSE [52] (Mini Mental State Examination; subsequently dropped from analysis) to enable translation of missingness structure from clinical data to ADNIMERGE data (see section II.B.2).

**TABLE I.**
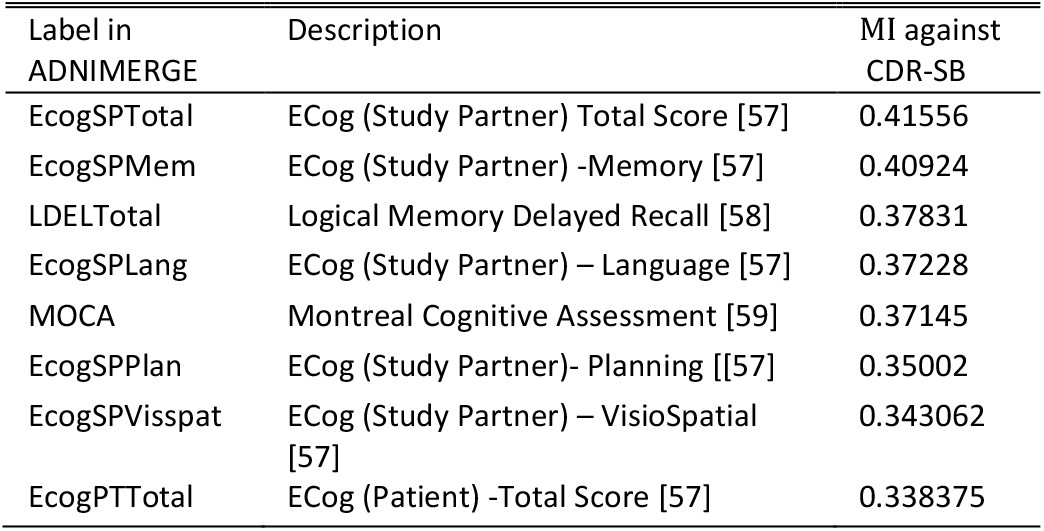
Features Selected By Mutual Information (MI) With Outcome

We made use of CDR-SB (Clinical Dementia Rating Sum of Boxes) instead of the more subjective clinical diagnosis [53]. CDR-SB was re-coded from the ADNIMERGE variable CDR (Clinical Dementia Rating) following the protocol in [54]. The mild, moderate and severe AD classes were amalgamated creating a three-class outcome variable: Healthy Controls (HC), MCI, and AD.

Importantly, we used the resulting ADNIMERGE data to: (i) create synthetic missing datasets from a complete ADNI dataset, based on the missingness structure of real-world clinical data as described in Section II.A.1; (ii) evaluate the various computational approaches; and (iii) develop our proposed workflow.

### B. Computational Methods

#### 1) Feature Selection

Feature selection was performed on the ADNIMERGE table using the mutual information (*MI*) algorithm [55]:

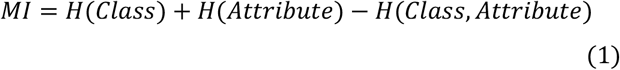

where H is Shannon’s entropy [56] defined by

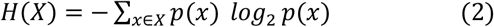

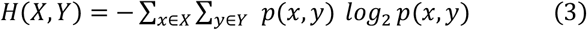

in which *P* is the probability function of some random variable *X* or *Y* for *p*ossible outcomes *x* and *y*, respectively. *H* can be understood as a measure of “disorder”: the sum of the probability of each label multiplied by the log probability of each label, with a value ranging between 0 and 1. The *MI* of a given attribute is the reduction in disorder of the class variable, when the class variable is separated according to that attribute.

The 8 CFAs which had the highest *MI* with respect to the CDR-SB outcome variable were selected. In addition, the MMSE score was retained to facilitate mapping of the types of missingness from the real-world clinical dataset, as described in Section II.B.2. Rows with original missing values for any of these features were dropped, creating an initial complete ADNIMERGE dataset with 1185 rows (the base dataset), with each row representing one individual participant visit. Multiple visits from the same participant at different time points were considered as separate cases here, as our original clinical data was not longitudinal. The dataset had imbalanced classes with 478 healthy controls, 614 MCI and 93 AD cases. This base dataset provided the ground truth for our study. Synthetic missing datasets were derived from this dataset for imputation and classification testing.

#### 2) Missing Data

Next, we searched for the relationship between missing values and the degree of cognitive decline of the individual/patient. Although no CFA in ADNIMERGE can be found in the clinical dataset, a previous study has provided a table of conversion between ACE-III scores (in our clinical dataset) and MMSE scores (in ADNIMERGE) [60]. In particular, these two CFAs were temporarily used to map the missingness structure from the clinical dataset to ADNIMERGE but subsequently not considered in the analysis (see below). We used the ACE-III scores in our clinical dataset as the benchmark for the relationship between missingness and cognitive decline, to facilitate this mapping without using the outcome variable for generating missingness (which would create double-dipping in subsequent analysis).

We first performed a regression of the proportion of missing values in the clinical dataset on ACE-III. The resultant regression equation (see Section III.1) was then used to generate synthetic missing data in the ADNIMERGE dataset. Specifically, the MMSE score in ADNIMERGE was converted into an ACE-III score using the conversion table in [60]. Missing values were then synthetically introduced into the CFA variables in the ADNIMERGE dataset using this conversion.

It should be noted that due to the different variables in the ADNIMERGE data compared to our real-world clinical data, no attempt was made to reproduce any column-wise missingness patterns from our clinical data, as this would not have reflected any true underlying relationships among variables in the new dataset. We showed, in Section III.A, that the proportion of missing data for CFA values was very high. Thus, in total, 10 synthetic ADNIMERGE datasets with different random missing patterns were generated, to ensure robustness in the results. ACE-III and MMSE scores were dropped from subsequent analysis, because ACE-III was not in ADNIMERGE and MMSE was not selected by feature selection.

#### 3) Data Imputation Methods

We included traditional mean and median data imputation methods [1] for analysis as they are straightforward to interpret and can function as a benchmark. We also used a multiple imputation method termed Predictive Mean Matching (PMM) [61]–[64] from the multivariate imputation via chained equations (MICE) package in R [65]. We used PMM both in the form of a single imputation (PMM1) and the mean of 5, 10, 15 and 50 imputations (PMM5, PMM10, PMM15 and PMM50, respectively). It should be noted that PMM is the default method for MICE, the most commonly used multiple imputation package. Imputation algorithms such as the k-NN method [35] which generalize from complete cases, were unsuitable for our high proportion of missing data, and were not considered.

The general steps for PMM within the context of MICE are as follows [64]: (i) linearly regress observed values for each column on the other columns, obtaining a set of coefficients; (ii) make a random draw from the posterior predictive distribution of this set of coefficients, creating a new set; (iii) use the newly generated coefficients to generate predictive values for missing values in this column (iv) identify a set of cases with observed variable whose predicted values are close to the predicted values for the case with missing data; and (v) from these cases, randomly choose one case and assign its observed value to substitute for the missing value. Steps (ii) to (v) are repeated for each column, and the whole process is iterated 10 times to generate one imputed dataset. For PMM1, one imputed dataset is generated, while for PMM5, 5 imputed datasets are generated (see Supplementary Fig. 1 for details).

Another algorithm which we used was the iterative missForest [18] from the missForest package in R [66], which uses Random Forest (RF) regression to impute missing data [67]. The missForest imputation method was chosen as it had been shown to outperform MICE at imputation [18], [68] and involved few assumptions about the structure of the missing data [18]. The MissForest method entails the following steps: (i) impute the column mean for each missing value in dataset D to create imputed dataset D’; (ii) copy D’ to D’’; (iii) for each column in D’ use the rows with no missing values to build a RF model, and use the model to predict the missing values; (iv) update D’ with new predictions for the missing values; (v) test convergence and output D’’ if convergence is reached – if maximum iterations have been reached output D’; otherwise iterate steps (ii-v) (see Supplementary Fig. 2 for details).

Finally, we also used the Bayesian Principal Components Analysis (BPCA) [69] algorithm for imputation as it has been found to be effective in previous studies [27], and in order to explore whether a PCA-based method impacts imputation accuracy by variable. Bayesian PCA is a computationally complex method which uses an iterative approach similar to Expectation Maximization, combined with Bayesian modelling to estimate the eigenvalues of the underlying principal components of the data (see Supplementary Fig. 3 for details).

The adjusted *R*^*2*^ of the linear regression of the imputed values on ground truth (complete data) was used as a measure of imputation accuracy, with values ranging from 0 to 1 (poorest to highest in accuracy, respectively). The mean, minimum and maximum *R*^*2*^ measurements from each of the 10 synthetic datasets were obtained. This methodology was also used to calculate the average imputation accuracy of each variable using the missForest algorithm.

The computation time over 10 missing datasets for each imputation method was recorded and normalized by dividing by the time for the fastest method (mean imputation)

#### 4) LOOCV Classification Accuracy Testing

Classification accuracy was tested using leave-one-out cross-validation (LOOCV) [70]. In the LOOCV condition, the test dataset is only one row. We used LOOCV to mimic one-patient classification condition. Further, LOOCV is suitable for smaller data sizes, which may occur in some clinical/medical centres. Although LOOCV is computationally intensive, it minimizes model bias by using almost all the training data for each classification while allowing conservative estimation [71]. The approaches we used for handling missing values in the test row can broadly be divided into two categories: 1) impute the missing values in the test row using the imputation approach used for the training dataset; or 2) use a reduced-feature classifier, where a classification model is built using only the features which are not missing in the test row. In a dataset with N rows, a classification model will be built N times and tested on each row in turn. A schematic of this process is shown in Fig. 3. Hyperparameter tuning using the bootstrap method with 3 repeats, and class balancing using downsampling, were incorporated within the “Build Classifier” step [68].

**Fig. 3.**
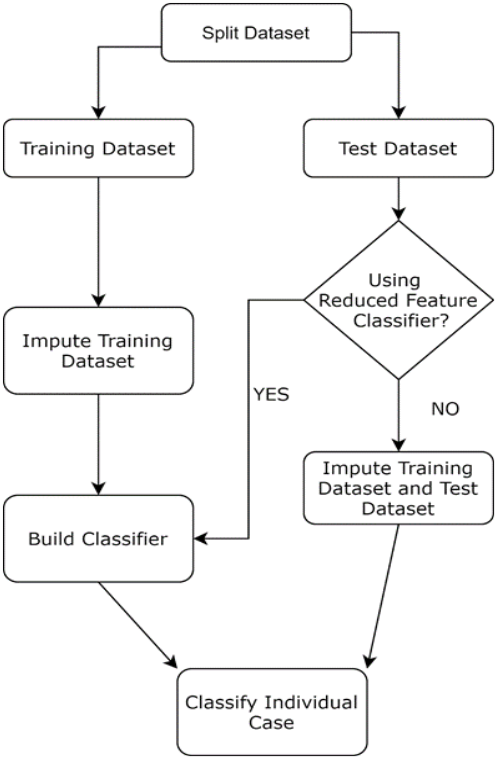
Workflow for LOOCV (single case) classification testing, emulating actual clinical decision-making conditions. Data is first split into training and single-case test datasets. Training and test datasets are imputed separately.

The workflows shown in Table II are different instantiations of the general workflow shown in Fig. 3 (except for workflow H where no imputation was used). The workflows consist of the combination of training dataset imputation method, test dataset imputation method, and classifier method. The RF classifier (from the caret R package [72]) was used in most cases, as it is versatile and adaptable to a wide variety of different datasets [18], with the SVM classifier (also from the caret package) used in some workflows to test whether imputation strategies have different compatibility with different classifiers. The naïve Bayes (NB) classifier (from the e1071 R package [73]) was used in (H) as it does not require a strategy for handling missing values; the classifier can skip a missing value while still making use of values in the same row of the dataset due to the conditional independence assumptions in the naïve Bayes algorithm. The RF imputation method was used as it was the most effective single imputation method, as well as multiple imputation with PMM-5 (higher values of multiple imputation were not considered here due to the impact on classification speed) and single imputation with the mean of PMM-15, which although not intended for single imputation was found to be both faster and more accurate as an imputation method than RF. Multiclass area-under-the-ROC curve, AUC [74], over the 1185 cases was calculated using the pROC package [75]. 95% AUC confidence intervals were bootstrapped with 500 resamples. We also provide in Supplementary Table I, sensitivity and specificity results, as well as a baseline comparison using RF, SVM and naïve Bayes classifiers on the complete dataset with no missing values. In a clinical decision support setting, imputation and classification will occur in different contexts, so the computation times for imputation and classification in each workflow were recorded separately. The mean computation time in seconds (s) for each of the 1185 classification and imputation cases was recorded.

**TABLE II.** Imputation and Classification Workflows

|  | Training dataset imputation | Test dataset treatment | Classifier | Imputation time (s) | Classification time (s) | AUC |
| --- | --- | --- | --- | --- | --- | --- |
| A | mean | mean | RF | 0.002 | 1.974 | 0.871 |
| B | class mean | reduced feature | RF | 0.010 | 1.297 | 0.878 |
| C | RF | RF | RF | 11.520 | 1.950 | 0.889 |
| D | mean | reduced feature | RF | 0.002 | 2.029 | 0.867 |
| E | RF | reduced feature | RF | 11.444 | 1.860 | 0.876 |
| F | PMM5 multiple | PMM5 multiple | modal imputed outcome | 3.179 | 0.021 | 0.839 |
| G | PMM5 multiple | PMM5 multiple | RF ensemble | 3.179 | 14.566 | 0.885 |
| H | none | none | NB | 0.000 | 0.000 | 0.885 |
| I | PMM5 multiple | reduced feature | RF ensemble | 3.179 | 10.100 | 0.891 |
| J | Mean | Mean | SVM | 0.002 | 1.717 | 0.867 |
| K | RF | RF | SVM | 11.520 | 2.313 | 0.882 |
| L | RF | reduced feature | SVM | 11.520 | 1.694 | 0.893 |
| M | PMM15 mean | RF | RF | 10.315 | 2.150 | 0.887 |
| N | PMM15 mean | Reduced feature | RF | 10.315 | 0.318 | 0.884 |
| O | PMM15 mean | RF | SVM | 10.315 | 0.122 | 0.888 |
| P | PMM15 mean | Reduced feature | SVM | 10.315 | 0.142 | 0.874 |

### C. Software and Hardware for Analysis

The above analyses and algorithms were run within R Studio version 1.146 on a Windows machine with eight memory cores, Intel i7 processor, 16GB Ram and R version 3.5.2 installed. The analyses were all single threaded to allow for straightforward comparison of computational cost. The codes are available at https://github.com/mac-n/BHI-missing-data.

## III. RESULTS

### A. Synthetic missing data with missingness type from clinical data

To reduce the size of the ADNIMERGE dataset to better resemble the real-world clinical dataset, we performed feature selection using the mutual information algorithm [55] which selected the best features with respect to the class variable (CDR-SB scores in our case), and identified the 8 most relevant CFA features. Table I shows the selected CFAs in descending order of their mutual information with the class variable. Interestingly, most of the selected CFAs were completed by study partners, who accompanied the patients to the study site throughout the ADNI study, as opposed to being completed by the patients themselves (Table I, column 2). Next, we used the top 8 CFAs, plus Gender and Age variables and our class variable from the ADNIMERGE data to form our baseline dataset which resembled the types of features in the memory clinic data. We then investigated the missingness in our memory clinic data, in order to reproduce the same missingness patterns in the ADNIMERGE data.

Using the memory clinic data, we determined that the data had MAR type missingness by regressing the number of missing values in each row, normalised by the number of CFA columns, on Addenbrooke’s Cognitive Examination (ACE-III.) The ACE scale was used because there is known mapping from ACE to MMSE scores [60]. Although there are no common CFAs between the memory clinic data and ADNIMERGE, MMSE scores are available in ADNIMERGE to recreate the same type of missingness in ADNIMERGE as found in our memory clinic data. Higher order fits were tested but higher order terms were found to be non-significant in the polynomial regression (2^nd^ order: *p* -value = 0.051; 3^rd^ order:*p* -value = 0.39).

We found that the resulting regression equation could be described by *N*_*miss*_ = *0*.*48 + (0*.*06 ACE-III)*, where *N*_*miss*_ was the proportion of CFA values missing in each row, and *ACE-III* consisted of its normalized score. The 0.48 constant in the equation meant that 48% of the CFA values were missing. The low *p-*value (*p* =2×10^−16^, *n*=189) and low *R*^*2*^ (0.02502) of the regression indicated that cognitive decline, as measured by ACE-III scores, was significantly correlated with missingness but cognitive decline could not explain most of the missingness in the data. Hence the data could be considered either MCAR or MAR. A conversion table to convert MMSE scores in ADNI to ACE-III scores [60] was used. The regression above was then used in combination with the generated ACE-III scores to generate a probability of missingness, *P*_*miss,i*_. for every row *i* in ADNIMERGE. Each variable in each row *i* was substituted with a missing value, with probability *P*_*miss,i*_. In this manner, 10 missing datasets were generated from the original complete ADNIMERGE data with the same degree and type of missingness as in our clinical data (see Section II.A.2).

### B. Computationally expensive imputation methods are not necessarily more accurate

Based on the synthetic missing datasets, we performed various imputation methods. We found that the Predictive Mean Matching (PMM) and Random Forest (RF) imputation methods provided the highest accuracy when tested against the complete dataset (ground truth) (Fig. 4). PMM imputation methods were further divided into PMM5, PMM10, PMM15, PMM50 - the mean of 5, 10, 15 and 50 multiple imputations, respectively. Specifically, the regression of the mean of the PMM50 imputation method against ground truth was the most accurate, with a mean *R*^*2*^ over 10 synthetic datasets of 0.86 (Fig. 4). This was non-significantly (*p*=0.204) higher than the accuracy when using PMM15 imputation (mean 0.861), but significantly higher than the accuracy for PMM10 (0.856) (t-test *p*-value over 10 datasets = 0.002). The PMM15 method was in turn significantly (*p*=0.001) more accurate than the RF method (mean 0.849) although the RF was the only method with accuracy close to the PMMs. Thus, PMM’s accuracy marginally increased when more multiple imputations were generated. All PMM methods involving more than 15 imputations were significantly more accurate than RF.

**Fig. 4.**
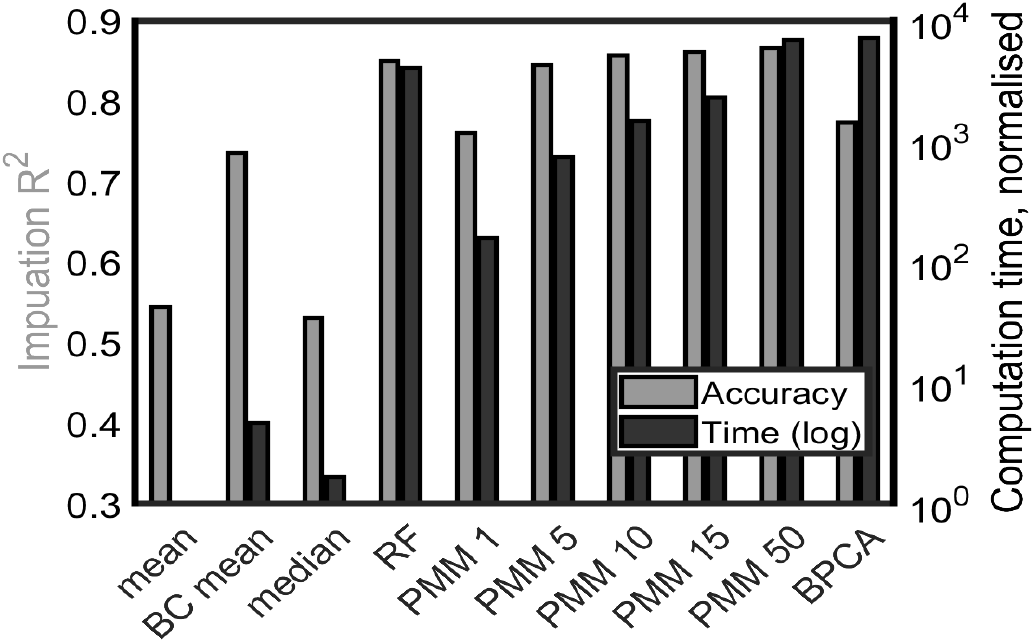
Imputation accuracy R^2^ and computation time depend on imputation methods. Grey (black) bars: accuracy R^2^ (computation time). Left-to-right bars: mean imputation, mean by class imputation, median imputation, RF imputation, PMM averaged over 1, 5, 10, 15 and 50 imputations, and BPCA.

The next most accurate method, Bayesian Principal Component Analysis (BPCA), was found to have an *R*^2^ of 0.773. The BC mean (mean by class) imputation method had a reasonable accuracy for a computationally simple method (*R*^*2*^=0.735), but as an imputation method it had the disadvantage that it could not be used to impute the test row as the class value of the test row was not known. Finally, the median and mean methods did not achieve high accuracy.

Given that many of the mean *R*^*2*^ values were between 0.8-0.9 (Fig. 4, grey bars), we next investigated the computational cost of individual imputation methods. We found that there was a wider range of computational times across the various imputation methods (Fig. 4, black bars; note the logarithmic scale). In particular, BPCA and PMM50 had similar timescales, while RF was about twice as fast. PMM15 was twice as fast as RF. The mean, BC mean and median methods, as might be expected, were not computationally costly. Overall, computationally expensive methods could achieve higher accuracy than simpler methods (e.g. RF and PMMs cf. mean, median and BC median), but algorithmic complexity did not guarantee high accuracy (e.g. BPCA).

### C. Running time varies logarithmically across workflows

Next, we investigated the most effective data imputation methods, with respect to classification accuracy and computational cost. We tested various workflows *A-P* (Table II; see Supplementary Table I for additional results) for classification and imputation of training and test datasets in the LOOCV condition, where each case in the dataset was classified one at a time, mimicking handling a single patient/individual (Section II.B.4). To demonstrate this, it sufficed to use just one of the synthetic datasets. The test dataset, consisting of only 1 row, was imputed either with the same imputation algorithm as the training dataset, or was not imputed and was classified using a reduced-feature classifier which used only features which were not missing in the test dataset. Class balancing and parameter tuning were incorporated within the classification step.

Among the workflows we tested, the multiclass AUC ranged between 0.83 and 0.89. This was a surprising result, given the extreme (48%) missingness that was introduced (and comparable to the AUCs using the complete dataset – see Supplementary Table I). Most of the workflows performed at similar levels, and the bootstrapped confidence intervals overlapped substantially. An outlier performing below the others was workflow F (AUC=0.839) which did not use a conventional classifier but a mode of multiple imputed values. Workflows J (mean imputation plus SVM classifier) and D (mean imputation + reduced feature random forest classifier) both performed relatively poorly with AUC below 0.87 - perhaps poor performance was unsurprising with simpler mean imputation, although it should be noted that mean imputation by class combined with a reduced feature random forest classifier (workflow B) performed better than many workflows deploying more sophisticated imputation methods.

We found workflows L (AUC=0.893), I (AUC=0.891), and C (AUC=0.889) to be ranked top in our results. Workflow L was a mixture of methods: RF imputation with a reduced feature SVM classifier. Workflow I used 5x multiple imputation and an ensemble of reduced feature RF classifiers, while workflow C imputed both training and test dataset with RF and used a RF classifier. Given that no particular approach substantially stood out in terms of AUC measure, we then investigated the computation time. In particular, the running time for the LOOCV workflows had been divided into classification time (time to build the classifier and perform classification) and imputation time (time to impute the training set) (presuming that in a clinical decision support setting, imputation was executed during off-peak times). Hence, workflows C, F, G and K, with test dataset imputed iteratively alongside the training dataset, might be impractical for use in a clinical decision-making setting if the dataset was large (Fig. 5) and we included them here primarily for benchmarking purposes. In terms of imputation time, RF imputation and PMM imputation methods were the slowest, and mean imputation methods were orders of magnitude faster.

**Fig. 5.**
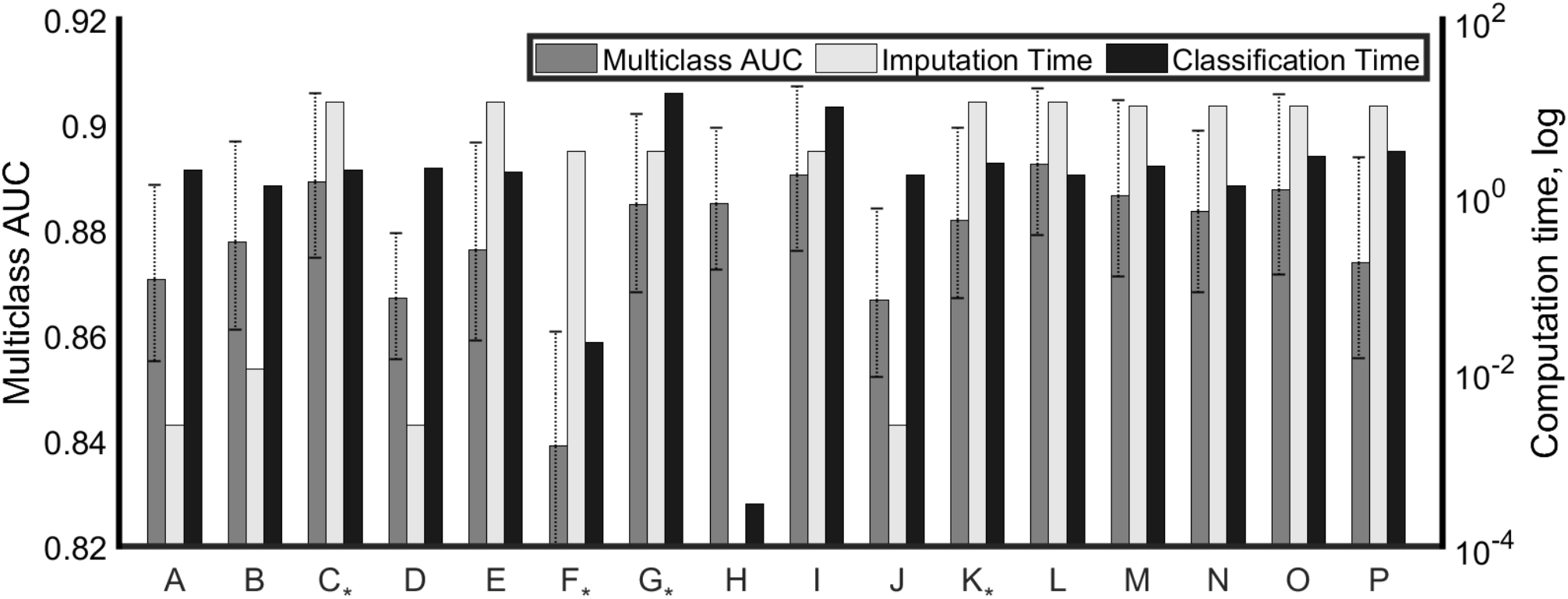
Imputation and classification workflows evaluated for multiclass AUC (light grey, left axis; linear scale), imputation time and classification time (respectively dark grey and black, right axis; logarithmic scale.) Details of the workflows are explained in Table 2. Workflows marked with * impute the test dataset alongside the training data; hence imputation and classification must be performed together.

An interesting outlier in terms of computation time was workflow H (naive Bayes classification), which used no imputation and was an exceptionally fast classifier with AUC=0.885. Other outliers in terms of classification time were workflows G and I which used an ensemble of RF classifiers and were hence considerably slower than other methods. In general, the reduced feature classifiers were faster to build than the classifiers which used all the features in the dataset.

## IV. DISCUSSION

Clinical datasets such as in electronic health records often have a significant proportion of missing data [3], [4], [6]. Various strategies have previously been proposed, however, there is no study that deals with the practical problem of missing dementia data in the test dataset, even though this is very likely to occur in clinical practice. This “test dataset” comprises the individual patient to be diagnosed. Missing data in the test dataset may prohibit the use of many popular imputation methods, which are frequently iterative and computationally costly when datasets are large. In this work, with a focus on AD diagnosis, we have replicated the missingness structure of a real-world routine (memory) clinical dataset and proposed practical strategies for dealing with a significant proportion (48%) of missing data in training and test datasets (Fig. 2). Moreover, we evaluated the approaches under the LOOCV condition (Fig. 1), mimicking real-world clinical decision-making (Fig. 3). We found that, despite the extreme missingness introduced, the AUC results from our proposed workflows were comparable to those produced using the original complete dataset (see Supplementary Table I).

Overall, we found that various strategies for imputation and classification in these conditions were able to maximise the classification AUC but these methods varied widely in computation time (Figs. 5 and 6), and this might likely be an important factor when developing or maintaining a clinical decision support system. In addition, an interesting finding from our feature selection was that partner evaluation was more informative regarding AD severity than self-evaluation, which may inform future design of dementia assessments.

In particular, reduced-feature methods for dealing with missing test datasets performed equally well to methods that involved imputing the test dataset, although this was sensitive to the imputation method. Reduced-feature methods might be the best solution for building a clinical decision support tool with large data as they did not involve real-time imputation of the test dataset. Specifically, we found RF imputation of the training dataset combined with a reduced-feature SVM classification (workflow L in Table II) was the best performing workflow and was also the fastest classifier to build. However, a drawback for reduced-feature methods is that either a large number of models must be stored, one for each possible combination of columns, or the classification model must be trained on-the-fly, and this will constitute part of the cost-benefit analysis when choosing a workflow for practical applications.

Mean imputation by class performed surprisingly well in our testing. This was despite the relatively low accuracy of mean imputation (Fig. 4) and was consistent with previous work suggesting that imputation accuracy did not always have a large effect on classification performance [76]. It could be argued that mean imputation is a form of missing-indicator imputation, as any missing value in a given column will have the same imputed value. Thus, the classification model receives a signal that the value was missing, which may improve classification in some circumstances – this will be explored in future work. When real-time computational speed is at a premium and the dataset has large number of features, mean imputation by class combined with a reduced-feature classifier (workflow B) may be worth investigating. However, the naïve Bayes classifier without imputation (workflow H) performed better than workflow B and had remarkably fast computation times. It may be the case that variants on the naïve Bayes approach, such as model averaged naïve Bayes [77], can provide an optimal solution in terms of both classification performance and computation time, especially when the number of features is large, and this will be investigated in future work.

Our present study has several limitations and could be extended in several ways. So far, we have only used one dataset from a memory clinic. In future studies, different clinical datasets with different types of clinical features will need to be explored to validate our results. Moreover, we have only investigated limited types of extreme missingness. Future work will investigate cases with less, and different types of missingness. This may involve more sophisticated models to generate complex missingness structures (e.g. column-wise missingness relationships). We have also not completely evaluated other imputation methods, such as those using unsupervised learning with autoencoders [31]. Their performance should be compared with the methods used in our current study. Further, this work has not completely explored the impact of relationships between features on the imputation process, which we have examined in more detail in [78].

In conclusion, we have suggested data imputation strategies for handling extreme missingness in both training and test data. Importantly, the strategies were proposed with practical applications in mind, especially for clinical decision support systems in dementia diagnosis. In terms of practical evaluation, we found that more complex and computationally costly methods did not offer significant advantage over more efficient methods.

## Data Availability

Data from the Alzheimer's Disease Neuroimaging Initiative (ADNI) is openly available.

https://github.com/mac-n/BHI-missing-data

## ACKNOWLEDGEMENTS

Data collection and sharing for this project was funded by the Alzheimer’s Disease Neuroimaging Initiative (ADNI) (National Institutes of Health Grant U01 AG024904) and DOD ADNI (Department of Defense award number W81XWH-12-2-0012). ADNI is funded by the National Institute on Aging, the National Institute of Biomedical Imaging and Bioengineering, and through generous contributions from the following: AbbVie, Alzheimer’s Association; Alzheimer’s Drug Discovery Foundation; Araclon Biotech; BioClinica, Inc.; Biogen; Bristol-Myers Squibb Company; CereSpir, Inc.; Cogstate; Eisai Inc.; Elan Pharmaceuticals, Inc.; Eli Lilly and Company; EuroImmun; F. Hoffmann-La Roche Ltd and its affiliated company Genentech, Inc.; Fujirebio; GE Healthcare; IXICO Ltd.; Janssen Alzheimer Immunotherapy Research & Development, LLC.; Johnson & Johnson Pharmaceutical Research & Development LLC.; Lumosity; Lundbeck; Merck & Co., Inc.; Meso Scale Diagnostics, LLC.; NeuroRx Research; Neurotrack Technologies; Novartis Pharmaceuticals Corporation; Pfizer Inc.; Piramal Imaging; Servier; Takeda Pharmaceutical Company; and Transition Therapeutics. The Canadian Institutes of Health Research is providing funds to support ADNI clinical sites in Canada. Private sector contributions are facilitated by the Foundation for the National Institutes of Health (www.fnih.org). The grantee organization is the Northern California Institute for Research and Education, and the study is coordinated by the Alzheimer’s Therapeutic Research Institute at the University of Southern California. ADNI data are disseminated by the Laboratory for Neuro Imaging at the University of Southern California.

See acknowledgements for ADNI below.

