## Supplementary materials for "Practical Strategies for Extreme Missing Data Imputation in Dementia Diagnosis"

### SUPPLEMENTARY TABLE I: IMPUTATION AND CLASSIFICATION WORKFLOWS ADDITIONAL RESULTS

| | Training dataset imputation | Test dataset treatment | Classifier | Imputa-tion time (s) | Classificat ion time (s) | AUC | Accuracy | Sensitivity (positive class MCI/AD) | Specificity (positive class MCI/AD) | Sensitivity (positive class AD) | Specificity (positive class AD) |
|---|---|---|---|---|---|---|---|---|---|---|---|
| A | mean | mean | RF | 0.002 | 1.974 | 0.871 | 0.680 | 0.786 | 0.824 | 0.839 | 0.878 |
| B | class mean | reduced feature | RF | 0.010 | 1.297 | 0.878 | 0.709 | 0.792 | 0.826 | 0.839 | 0.904 |
| C | RF | RF | RF | 11.520 | 1.950 | 0.889 | 0.743 | 0.864 | 0.795 | 0.828 | 0.913 |
| D | mean | reduced feature | RF | 0.002 | 2.029 | 0.867 | 0.632 | 0.767 | 0.818 | 0.914 | 0.831 |
| E | RF | reduced feature | RF | 11.444 | 1.860 | 0.876 | 0.716 | 0.839 | 0.789 | 0.839 | 0.896 |
| F | PMM5 multiple | PMM5 multiple | modal imputed outcome | 3.179 | 0.021 | 0.839 | 0.740 | 0.833 | 0.743 | 0.548 | 0.976 |
| G | PMM5 multiple | PMM5 multiple | RF ensemble | 3.179 | 14.566 | 0.885 | 0.738 | 0.851 | 0.797 | 0.806 | 0.918 |
| H | none | none | NB | 0.000 | 0.003 | 0.885 | 0.713 | 0.699 | 0.900 | 0.796 | 0.945 |
| I | PMM5 multiple | reduced feature | RF ensemble | 3.179 | 10.100 | 0.891 | 0.736 | 0.836 | 0.822 | 0.860 | 0.905 |
| J | Mean | Mean | SVM | 0.002 | 1.717 | 0.867 | 0.674 | 0.784 | 0.774 | 0.806 | 0.902 |
| K | RF | RF | SVM | 11.520 | 2.313 | 0.882 | 0.729 | 0.849 | 0.789 | 0.806 | 0.913 |
| L | RF | reduced feature | SVM | 11.520 | 1.694 | 0.893 | 0.727 | 0.823 | 0.845 | 0.871 | 0.895 |
| M | PMM15 mean | RF | RF | 10.315 | 2.150 | 0.887 | 0.751 | 0.859 | 0.799 | 0.796 | 0.927 |
| N | PMM15 mean | Reduced feature | RF | 10.315 | 0.318 | 0.884 | 0.712 | 0.843 | 0.772 | 0.860 | 0.899 |
| O | PMM15 mean | RF | SVM | 10.315 | 0.122 | 0.888 | 0.749 | 0.868 | 0.787 | 0.817 | 0.920 |
| P | PMM15 mean | Reduced feature | SVM | 10.315 | 0.142 | 0.874 | 0.714 | 0.857 | 0.743 | 0.828 | 0.907 |
| | | | | | | *Ground truth with no missing data* | | | | | |
| - | - | - | RF | - | 0.862 | 0.9101 | 0.784 | 0.883 | 0.851 | 0.871 | 0.918 |
| - | - | - | SVM | - | 0.125 | 0.9119 | 0.783 | 0.874 | 0.864 | 0.903 | 0.913 |
| - | - | - | NB | - | 0.007 | 0.8944 | 0.722 | 0.706 | 0.939 | 0.838 | 0.929 |

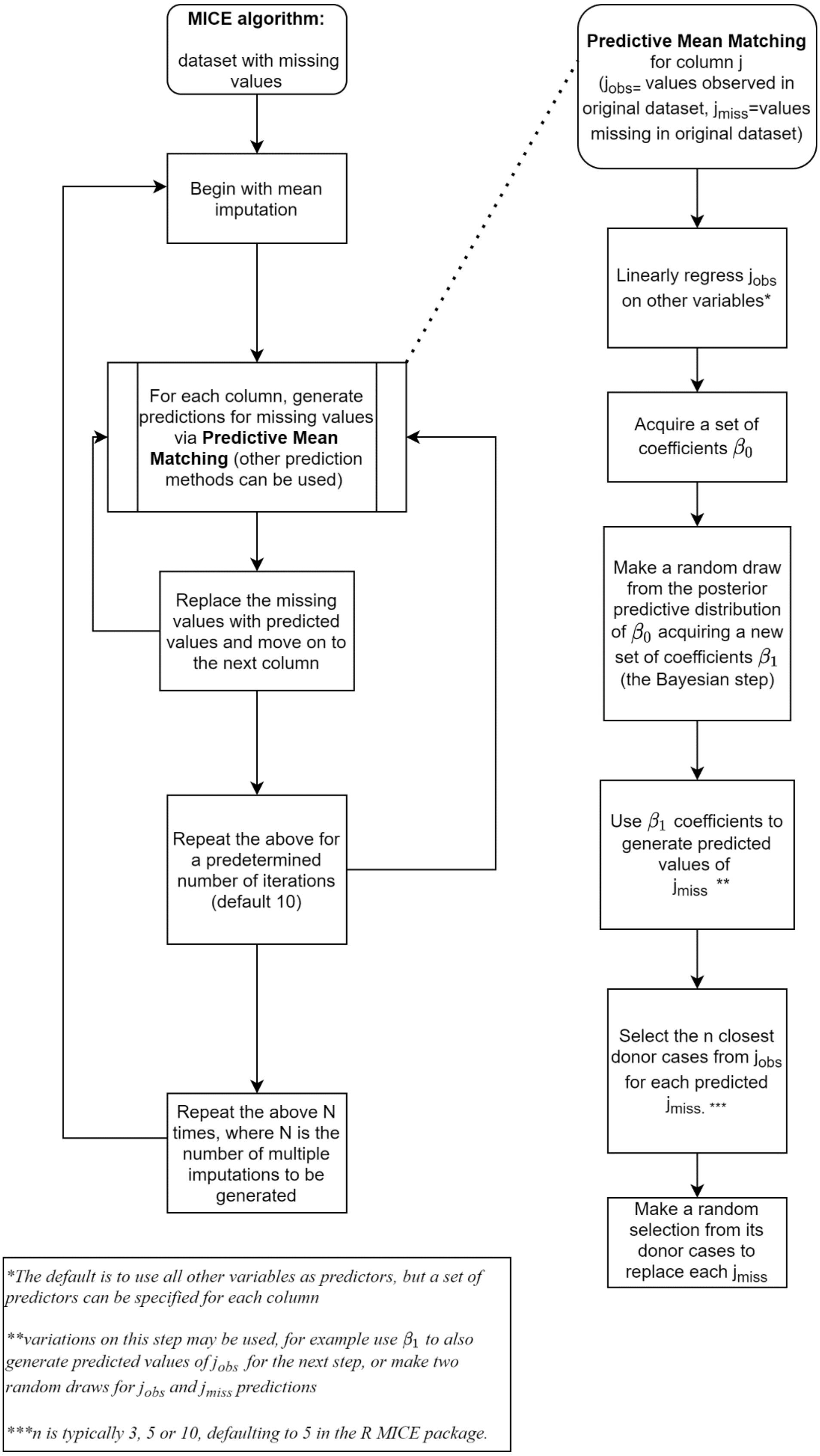


Supplementary Fig. 1. Multiple Imputation by Chained Equations (MICE) using Predictive Mean Matching (PMM). See [1]–[3] for further details.

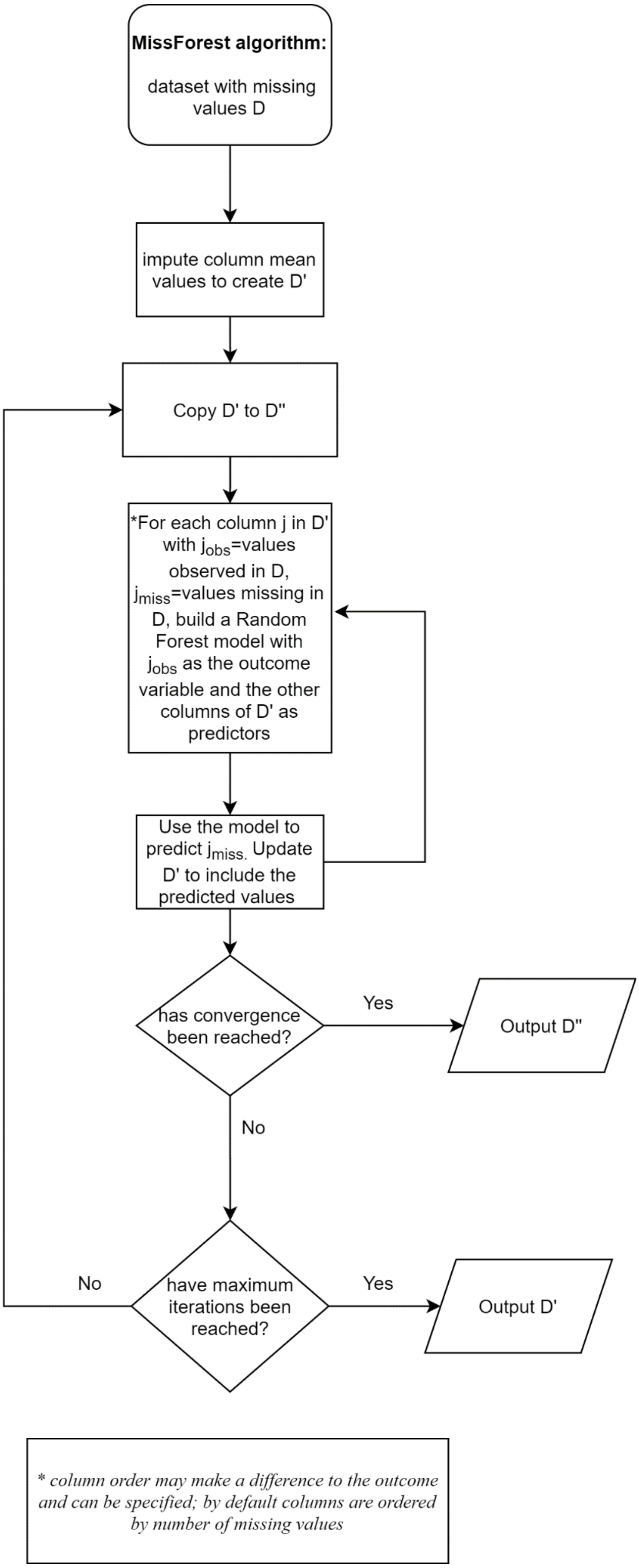


Supplementary Fig. 2. MissForest imputation algorithm. See [4] for further details.

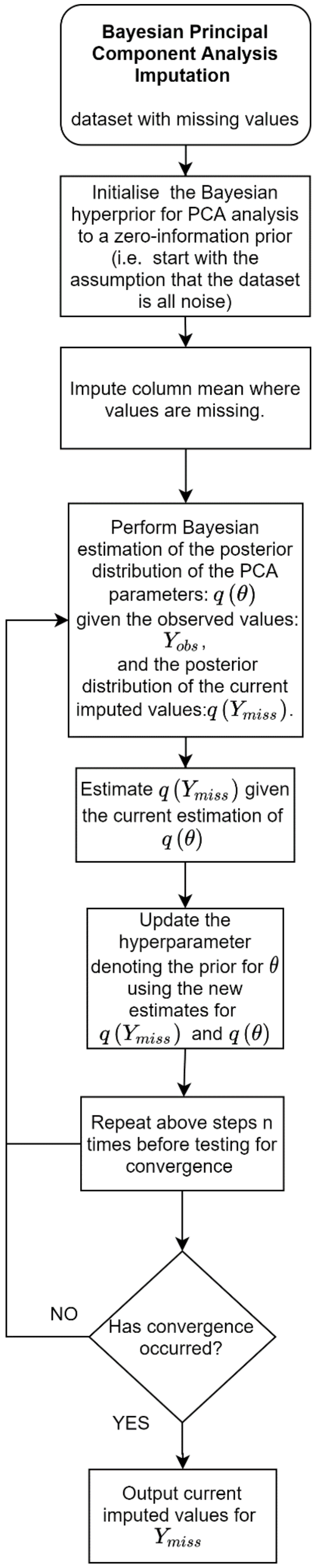


Supplementary Fig. 3. Imputation by Bayesian Principal Component Analysis (BPCA). The goal is to estimate the weights, loadings and noise (denote collectively as $\theta$) of a principal component analysis (PCA) of the data in the presence of missing values. Simultaneously, use the estimated PCA, $\theta$, to generate imputed values for the values that are missing -$Y_{miss}$.. An iterative estimation method is used, where $Y_{miss}$ is imputed first, then $\theta$ is estimated, followed by imputation of $Y_{miss}$, etc. This series of alternating iterative steps is similar to the well-known Expectation Maximisation algorithm, but here the computation is on the posterior distribution (i.e. likelihood adjusted for prior) of $\theta$, $q(\theta)$, and the posterior distribution of $Y_{miss}$, $q(Y_{miss,})$, rather than the likelihood. This method of missing value imputation is developed in [5], which is based on prior work in [6] and [7]. The version used in the paper was implemented in the pcaMethods package [8] in R [9].

[9] R Core Team, “R: A Language and Environment for Statistical Computing.” Vienna, Austria, 2019.
